# Perioperative diabetes related complications in patients undergoing major surgery are associated with the duration of gastrointestinal and orthopaedic surgery

**DOI:** 10.64898/2026.09.14.26362996

**Authors:** Cyril J Chacko, Orieanna Reeve Chen, Arundhati Binuraj, Sanjana Cherukad, Umar Saifuddin, Shirin Nourolahi-Oskoui, Amy Nyarko, Zubair Ahmed, Subashini Suresh, Suresh Renugappa, Deepak Singh Ranger, Rajeev Raghavan, Prashant Nasa, Tonny Veenith

## Abstract

**Background:** Diabetes mellitus (DM) is a common chronic condition among surgical patients. It is associated with increased perioperative morbidity and mortality. Real-world data on the clinical predictors of postoperative complications in diabetic patients undergoing major surgery are limited. This study aimed to characterise perioperative outcomes and identify independent predictors of 28-day postoperative complications.

**Methods:** A retrospective cohort study at a single UK tertiary hospital. Adult patients with a known diagnosis of type 1 or type 2 diabetes mellitus who underwent major procedures under general anaesthesia between January 2020 and September 2025 were identified through electronic patient records (EPR). The primary outcome was any postoperative complication within 28 days. We used multivariable logistic regression to identify independent predictors of complications, adjusting for age, sex, ASA grade, surgical category, admission urgency, operative duration, diabetes type, HbA1c, and socioeconomic deprivation (Index of Multiple Deprivation [IMD]).

**Results:** We screened 2623 patients and included 267; we analysed 267 patients (260 with outcome data). The 28-day postoperative complication rate was 23.1% (46/199) among those with outcome data. Infection was the most common complication. In multivariate analysis, surgical duration was the only independent predictor of complications (adjusted odds ratio [aOR], 1.71; 95% CI, 1.16–2.51; p=0.007). Other variables were not associated with complications.

**Conclusions:** In this study of patients with DM undergoing major surgery, operative duration was the only modifiable independent predictor of 28-day postoperative complications. Socioeconomic deprivation and preoperative HbA1c were not independently associated with outcomes, suggesting that standardised perioperative care pathways may attenuate these effects.

## Introduction

Diabetes mellitus (DM) is the most prevalent chronic condition in surgical practice. Over 5 million people in the United Kingdom live with DM, and the condition affects 15–18% of all hospital inpatients, roughly three times the background population prevalence^1,2^. Up to half of people with DM will require at least one surgical procedure during their lifetime, and it is estimated that more than 323,000 surgical procedures are performed annually on UK patients with diabetes^3^. The management of this growing population places substantial demands on anaesthetic and surgical services. Patients with diabetes carry a significantly higher burden of perioperative risk than the general surgical population. The surgical stress response produces counter-regulatory hyperglycaemia through catecholamines, cortisol, and glucagon, which is exaggerated and prolonged in patients with pre-existing insulin resistance or deficiency. Perioperative hyperglycaemia impairs neutrophil function, reduces tissue oxygen delivery, and disrupts wound healing, all of which contribute to elevated rates of surgical site infection, anastomotic leakage, acute kidney injury, and cardiovascular events^4,5^. A meta-analysis of 125 studies encompassing over 3.2 million patients reported an adjusted odds ratio of 1.65 (95% CI, 1.49–1.84) for any postoperative complication among patients with diabetes undergoing non-cardiac surgery^6^. Perioperative mortality may be up to 50% greater in patients with diabetes than in non-diabetic patients^7^.

National guidelines from the Joint British Diabetes Societies for Inpatient Care (JBDS-IP), the Association of Anaesthetists (2015), and the Centre for Perioperative Care (CPOC,updated2023) recommend a target capillary blood glucose of 6–10 mmol/L, preoperative HbA1c measurement within three months of elective surgery, and an HbA1c threshold of <69 mmol/mol for elective procedures^8,9^. Despite this report, the 2018 National Confidential Enquiry into Patient Outcome and Death (NCEPOD) report, Highs and Lows, identified suboptimal care in 36% of reviewed cases and absence of a perioperative diabetes management plan in nearly half of patients^10^.

Prospective data linking real-world perioperative practice to clinical outcomes in contemporary UK diabetic surgical patients remains limited. In particular, the relative contributions of operative duration, glycaemic control, and socioeconomic deprivation to postoperative outcomes have not been well characterised. This study aimed to describe the postoperative complication profile of adults with DM undergoing major surgery at a UK tertiary centre and to identify independent clinical predictors of 28-day postoperative complications using multivariable regression.

## Methods

A retrospective cohort study was conducted at a single UK tertiary hospital. The study was conducted and reported in accordance with the Strengthening the Reporting of Observational Studies in Epidemiology (STROBE). Data were extracted from the hospital electronic patient record (EPR) system.

### Eligibility Criteria

All consecutive patients with a diagnosis of Diabetes Meillitus who underwent major surgery between October 2020 and September 2025 were screened Patients were eligible for inclusion if they: (i) were aged 18 years or over; (ii) had a pre-existing documented diagnosis of type 1 or type 2 diabetes mellitus; and (iii) underwent major surgery under general anaesthesia, defined as joint replacement surgery, body cavity surgery (abdominal or pelvic), or other procedures requiring general anaesthetic with an operative duration exceeding 20 minutes. Patients were excluded if they underwent: peripheral limb surgery (non-joint replacement); day-case procedures; operations with a theatre duration of 20 minutes or less; obstetric surgery; or if they had newly diagnosed perioperative hyperglycaemia without a prior diagnosis of diabetes mellitus.extracted variable included: demographics (age, sex); clinical status (ASA physical status grade, frailty score where documented); diabetes-specific variables (type, HbA1c within three months of surgery, antidiabetic medications on admission and discharge); surgical details (procedure type, operative theatre duration, admission urgency, surgical specialty); perioperative outcomes (28-day complications, mortality, readmission); length of hospital stay; and patient postcode, used to derive socioeconomic status. Using validated Index of Multiple Deprivation (IMD), categorised as high deprivation (IMD deciles 1–3), intermediate deprivation (deciles 4–7), and low deprivation (deciles 8–10).

### Outcome Definitions

The primary outcome was any postoperative complication occurring within 28 days of surgery. Complications were recorded across pre-specified categories: infective (surgical site infection [SSI], sepsis, urinary tract infection), renal (acute kidney injury [AKI]), respiratory (pneumonia, respiratory failure), cardiac (arrhythmia, prolonged hypotension), haematological (anaemia requiring transfusion), endocrine (diabetic ketoacidosis, severe hypoglycaemia), neurological, and hepatic. Complication severity was graded using the Clavien-Dindo classification. Secondary outcomes included: major complications (Clavien-Dindo grade III–V); 28-day mortality; 28-day hospital readmission; ICU/HDU admission; and total length of hospital stay.

### Statistical Analysis

Continuous variables were compared between the complication and no-complication groups using the Mann-Whitney U test (normality assessed by the Shapiro-Wilk test). Categorical variables were compared using the chi-square test or Fisher’s exact test. Complication rates were compared across IMD deprivation groups using chisquare testing. Three multivariable logistic regression models were constructed to identify independent predictors of postoperative complications. Model 1A included age, sex, ASA grade, IMD group, surgical category, admission urgency, and operative duration. Model 1B additionally included diabetes type and HbA1c category (≥69 mmol/mol). Model 2 specifically examined the adjusted association between IMD deprivation and complications. Continuous variables (age, operative duration) were standardised prior to entry (z-score). Adjusted odds ratios (aOR) with 95% confidence intervals are reported. A complete case analysis was used throughout. Missing data were assessed descriptively. The frailty score was excluded from the primary regression models due to its high proportion of missing values (48.3%), reflecting protocol-mandated recording only for patients aged ≥65 years; it was retained in the univariable analysis. A two-sided p-value of <0.05 was considered statistically significant.

### Ethical Considerations

As a retrospective analysis of routinely collected EPR data with no patient identifiers retained in the analysis dataset, formal research ethics committee review was not required under UK Health Research Authority guidance. Institutional approval was obtained from the host university (SOABE/202526/staff/6) prior to data extraction. The study complied with the UK GDPR and the Data Protection Act 2018.

## Results

Of 2,623 patients screened from the EPR, 2356 were excluded (predominantly local anaesthetic, ophthalmological, minor orthopaedic and obstetric procedures), leaving 267 records (260 unique patients) for final analysis (Figure 1). The cohort was predominantly female (56.9%, n=152) with a median age of 62.0 years (IQR 45.5–71.0, range 17–91). Type 2 DM was present in 61.8% and type 1 DM in 38.2% of those with a recorded diabetes type (n=246). Most patients were ASA grade 3 (62.1%), and 69.0% were emergency admissions. Orthopaedic procedures (joint replacements and fracture fixation) accounted for 45.3% of cases, abdominal and other body cavity surgery for 44.9%, and cardiothoracic surgery for 9.7%. The cohort was disproportionately drawn from deprived areas, with 58.1% residing in IMD deciles 1–3 (high deprivation). Median preoperative HbA1c was 62.0 mmol/mol (IQR 50.8–75.0), and 34.9% had an HbA1c ≥69 mmol/mol at the time of surgery. Median operative theatre duration was 154.0 minutes (IQR 109.5–217.5). Baseline characteristics are summarised in Table 1.

**Table 1.**
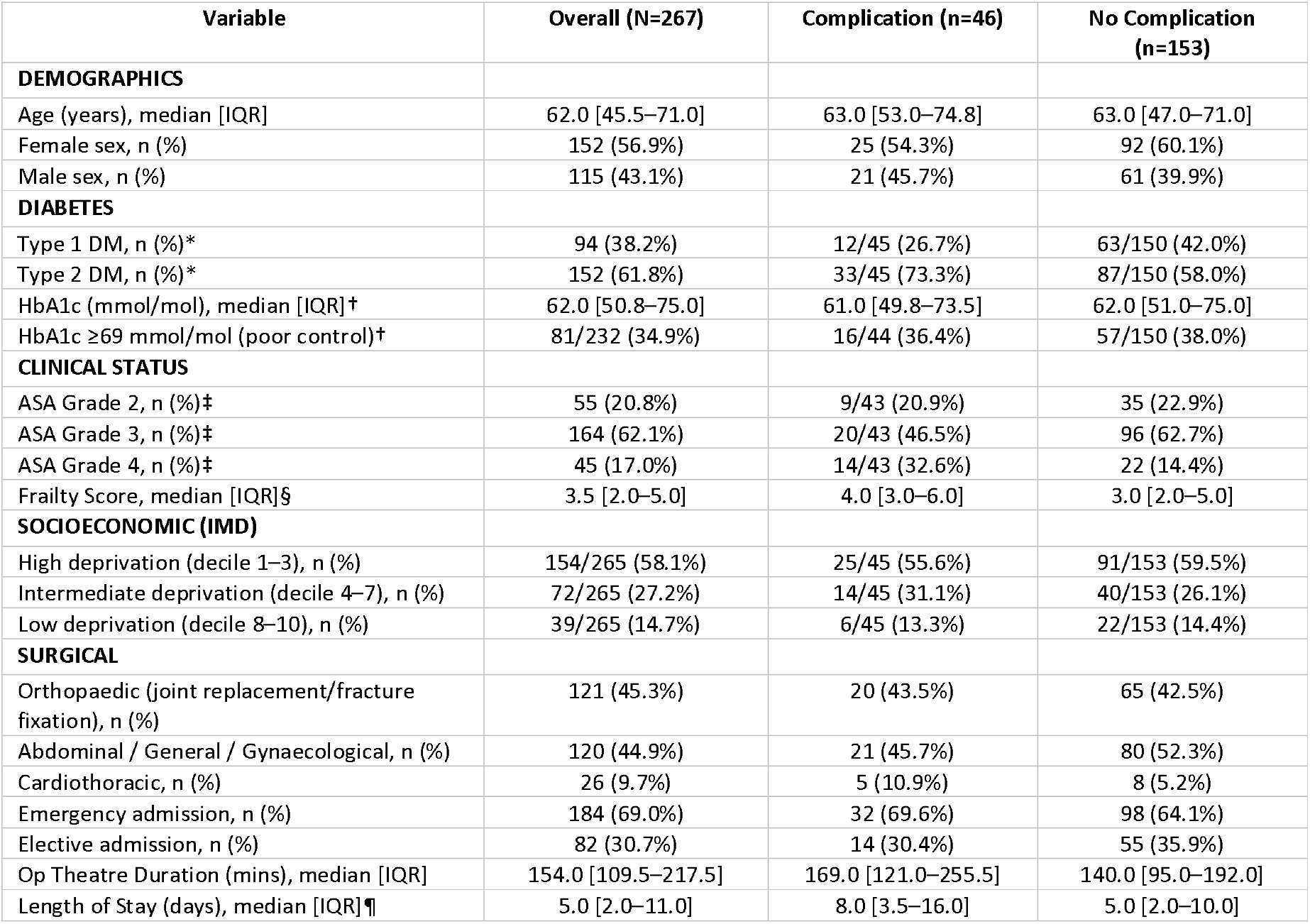
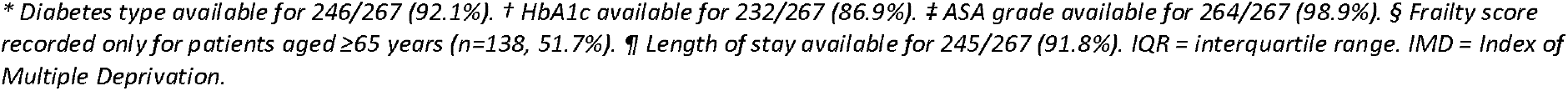
Baseline Characteristics: Overall Cohort and by 28-Day Complications.

| Variable | Overall (N=267) | Complication (n=46) | No Complication (n=153) |
| --- | --- | --- | --- |
| **DEMOGRAPHICS** |  |  |  |
| Age (years), median [IQR] | 62.0 [45.5–71.0] | 63.0 [53.0–74.8] | 63.0 [47.0–71.0] |
| Female sex, n (%) | 152 (56.9%) | 25 (54.3%) | 92 (60.1%) |
| Male sex, n (%) | 115 (43.1%) | 21 (45.7%) | 61 (39.9%) |
| **DIABETES** |  |  |  |
| Type 1 DM, n (%)* | 94 (38.2%) | 12/45 (26.7%) | 63/150 (42.0%) |
| Type 2 DM, n (%)* | 152 (61.8%) | 33/45 (73.3%) | 87/150 (58.0%) |
| HbA1c (mmol/mol), median [IQR]† | 62.0 [50.8–75.0] | 61.0 [49.8–73.5] | 62.0 [51.0–75.0] |
| HbA1c ≥69 mmol/mol (poor control)† | 81/232 (34.9%) | 16/44 (36.4%) | 57/150 (38.0%) |
| **CLINICAL STATUS** |  |  |  |
| ASA Grade 2, n (%)‡ | 55 (20.8%) | 9/43 (20.9%) | 35 (22.9%) |
| ASA Grade 3, n (%)‡ | 164 (62.1%) | 20/43 (46.5%) | 96 (62.7%) |
| ASA Grade 4, n (%)‡ | 45 (17.0%) | 14/43 (32.6%) | 22 (14.4%) |
| Frailty Score, median [IQR]§ | 3.5 [2.0–5.0] | 4.0 [3.0–6.0] | 3.0 [2.0–5.0] |
| **SOCIOECONOMIC (IMD)** |  |  |  |
| High deprivation (decile 1–3), n (%) | 154/265 (58.1%) | 25/45 (55.6%) | 91/153 (59.5%) |
| Intermediate deprivation (decile 4–7), n (%) | 72/265 (27.2%) | 14/45 (31.1%) | 40/153 (26.1%) |
| Low deprivation (decile 8–10), n (%) | 39/265 (14.7%) | 6/45 (13.3%) | 22/153 (14.4%) |
| **SURGICAL** |  |  |  |
| Orthopaedic (joint replacement/fracture fixation), n (%) | 121 (45.3%) | 20 (43.5%) | 65 (42.5%) |
| Abdominal / General / Gynaecological, n (%) | 120 (44.9%) | 21 (45.7%) | 80 (52.3%) |
| Cardiothoracic, n (%) | 26 (9.7%) | 5 (10.9%) | 8 (5.2%) |
| Emergency admission, n (%) | 184 (69.0%) | 32 (69.6%) | 98 (64.1%) |
| Elective admission, n (%) | 82 (30.7%) | 14 (30.4%) | 55 (35.9%) |
| Op Theatre Duration (mins), median [IQR] | 154.0 [109.5–217.5] | 169.0 [121.0–255.5] | 140.0 [95.0–192.0] |
| Length of Stay (days), median [IQR]¶ | 5.0 [2.0–11.0] | 8.0 [3.5–16.0] | 5.0 [2.0–10.0] |

\* Diabetes type available for 246/267 (92.1%). † HbA1c available for 232/267 (86.9%). ‡ ASA grade available for 264/267 (98.9%). § Frailty score recorded only for patients aged ≥65 years (n=138, 51.7%). ¶ Length of stay available for 245/267 (91.8%). IQR = interquartile range. IMD = Index of Multiple Deprivation.

**Figure 1.**
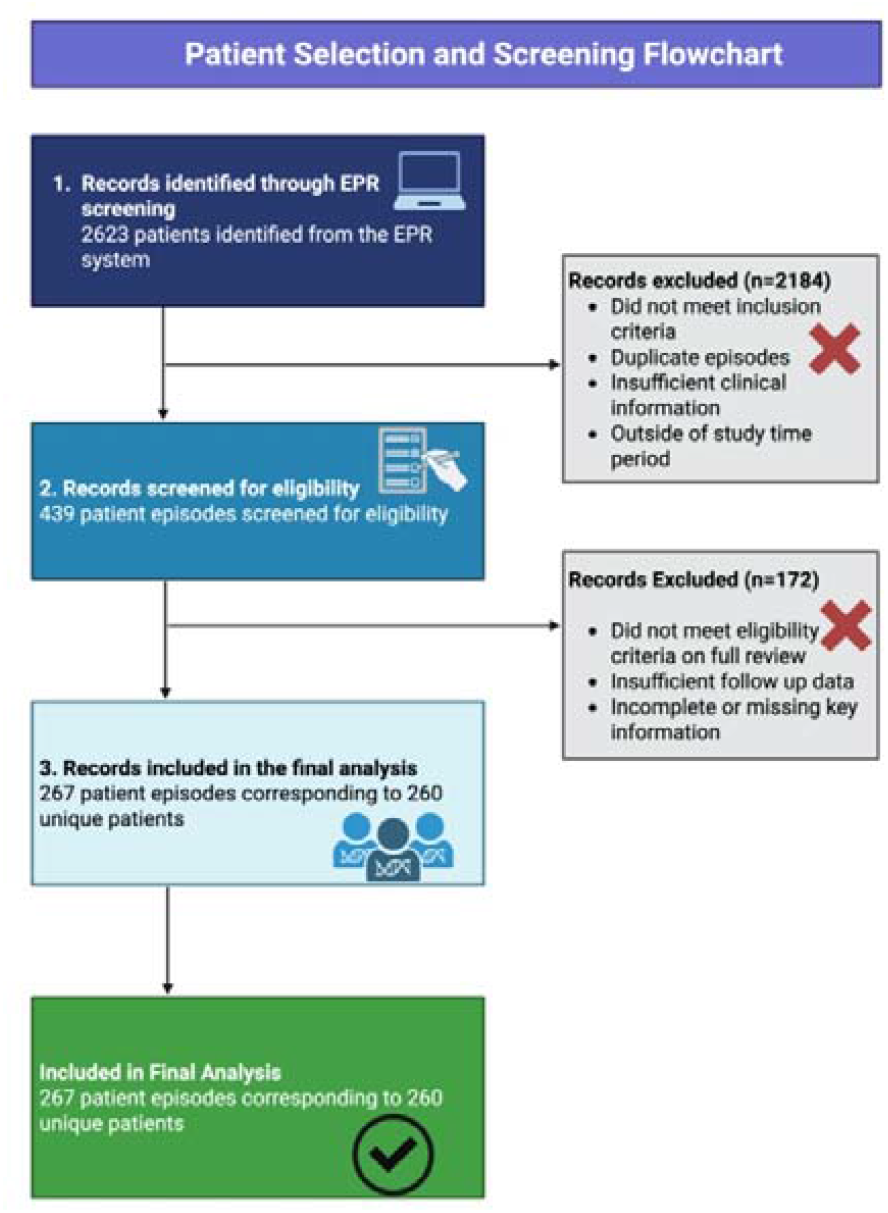
Patient selection and Screening flowchart. Created in https://BioRender.com

Complication outcome data were available for 199 of 267 records (74.5%). The 28-day complication rate was 23.1%(46/199). 28-day mortality was 2.1% (5/239 with mortality data) and readmission was 8.1% (19/235 with readmission data). Of 48 patients with Clavien-Dindo grading, 34 (70.8%) had minor complications (grades I–II) and 14 (29.2%) had major complications (grades III–V), including 6 in-hospital deaths (grade V). Outcomes are summarised in Table 2.

**Table 2.** Primary and Secondary Outcomes.

| Outcome | Events (n) | Denominator | Rate |
| --- | --- | --- | --- |
| 28-day complications (any) | 46 | 199 (with outcome data) | 23.1% |
| Major complications (Clavien-Dindo III–V) | 14 | 48 (with C-D grading) | 29.2% |
| 28-day mortality | 5 | 239 (with outcome data) | 2.1% |
| 28-day readmission | 19 | 235 (with outcome data) | 8.1% |
| ICU/HDU admission (Clavien IVa/b) | 4 | 48 (with C-D grading) | 8.3% |
| Clavien-Dindo Grade I | 17 | 48 | 35.4% |
| Clavien-Dindo Grade II | 17 | 48 | 35.4% |
| Clavien-Dindo Grade IIIa–b | 4 | 48 | 8.3% |
| Clavien-Dindo Grade IVa–b | 4 | 48 | 8.3% |
| Clavien-Dindo Grade V (death in hospital) | 6 | 48 | 12.5% |

### Complications

Infection was a frequent complication category, accounting for 31 events. Sepsis(n=13) was the most common specific infective complication, followed by urinary tract infection (n=5), and surgical site infections of various depths (organ space n=4, superficial n=4, deep n=4). Acute kidney injury was the most common non-infective complication (n=15). Followed by respiratory complications occurred in 13 cases (postoperative pneumonia n=8, respiratory failure n=5), cardiac complications in 11 (prolonged hypotension n=6, arrhythmia n=5), haematological complications in 9 (all anaemia requiring transfusion), and endocrine complications in 5 (DKA n=3, severe hypoglycaemia n=2). Six patients developed neurological or hepatic complications

### Univariate analysis

three variables differed significantly between the complication and no-complication groups: operative theatre duration (median 169.0 vs 140.0 mins; p=0.014), length of hospital stay (median 8.0 vs 5.0 days; p=0.028), and frailty score (median 4.0 vs 3.0; p=0.046), noting that frailty data were available in 138 patients only. ASA grade distribution also differed significantly (p=0.022), with 32.6% of patients in the complication group being ASA grade 4, compared to 14.4% in the no-complication group. Age, sex, HbA1c, diabetes type, admission urgency, surgical category, and IMD deprivation group did not differ significantly between groups Table 3. Complication rates were comparable across deprivation groups: 21.6% (high), 25.9% (intermediate), and 21.4%(low), P=0.805

**Table 3.** Univariable Comparison: Complication vs No-Complication Group.

| Variable | Complication (n=46) | No Complication (n=153) | p-value |
| --- | --- | --- | --- |
| Age (years), median [IQR] | 63.0 [53.0–74.8] | 63.0 [47.0–71.0] | 0.241 |
| Op Duration (mins), median [IQR] | 169.0 [121.0–255.5] | 140.0 [95.0–192.0] | **0.014 \*** |
| HbA1c (mmol/mol), median [IQR] | 61.0 [49.8–73.5] | 62.0 [51.0–75.0] | 0.380 |
| Length of Stay (days), median [IQR] | 8.0 [3.5–16.0] | 5.0 [2.0–10.0] | **0.028 \*** |
| Frailty Score, median [IQR] | 4.0 [3.0–6.0] | 3.0 [2.0–5.0] | **0.046 \*** |
| IMD Decile, median [IQR] | 3.0 [2.0–5.0] | 3.0 [2.0–5.0] | 0.793 |
| Sex (Male), n (%) | 21 (45.7%) | 61 (39.9%) | 0.598 |
| ASA Grade distribution | See Table 1 | See Table 1 | **0.022 \*** |
| Diabetes Type 2, n (%) | 33/45 (73.3%) | 87/150 (58.0%) | 0.093 |
| IMD deprivation group | — | — | 0.805 |
| HbA1c ≥69 mmol/mol, n (%) | 16/44 (36.4%) | 57/150 (38.0%) | 0.984 |
| Emergency admission, n (%) | 32 (69.6%) | 98 (64.1%) | 0.609 |
| Surgical category | — | — | 0.361 |

\* p<0.05. MWU = Mann-Whitney U test. All continuous variables were non-normally distributed (Shapiro-Wilk p<0.05). Denominators per variable reflect available data.

### Multivariate regression

Results are presented in Table 4 and Figure 2. In Model 1A (n=196 complete cases, 43 events), operative theatre duration was the only variable independently associated with postoperative complications (aOR 1.71 per standard deviation increase in duration, 95% CI 1.16–2.51, p=0.007). ASA grade 4 showed a clinically plausible but non-significant association compared with ASA grade 2 (aOR 2.24, 95% CI 0.74–6.78, p=0.154). No other variable reached statistical significance.

**Table 4.**
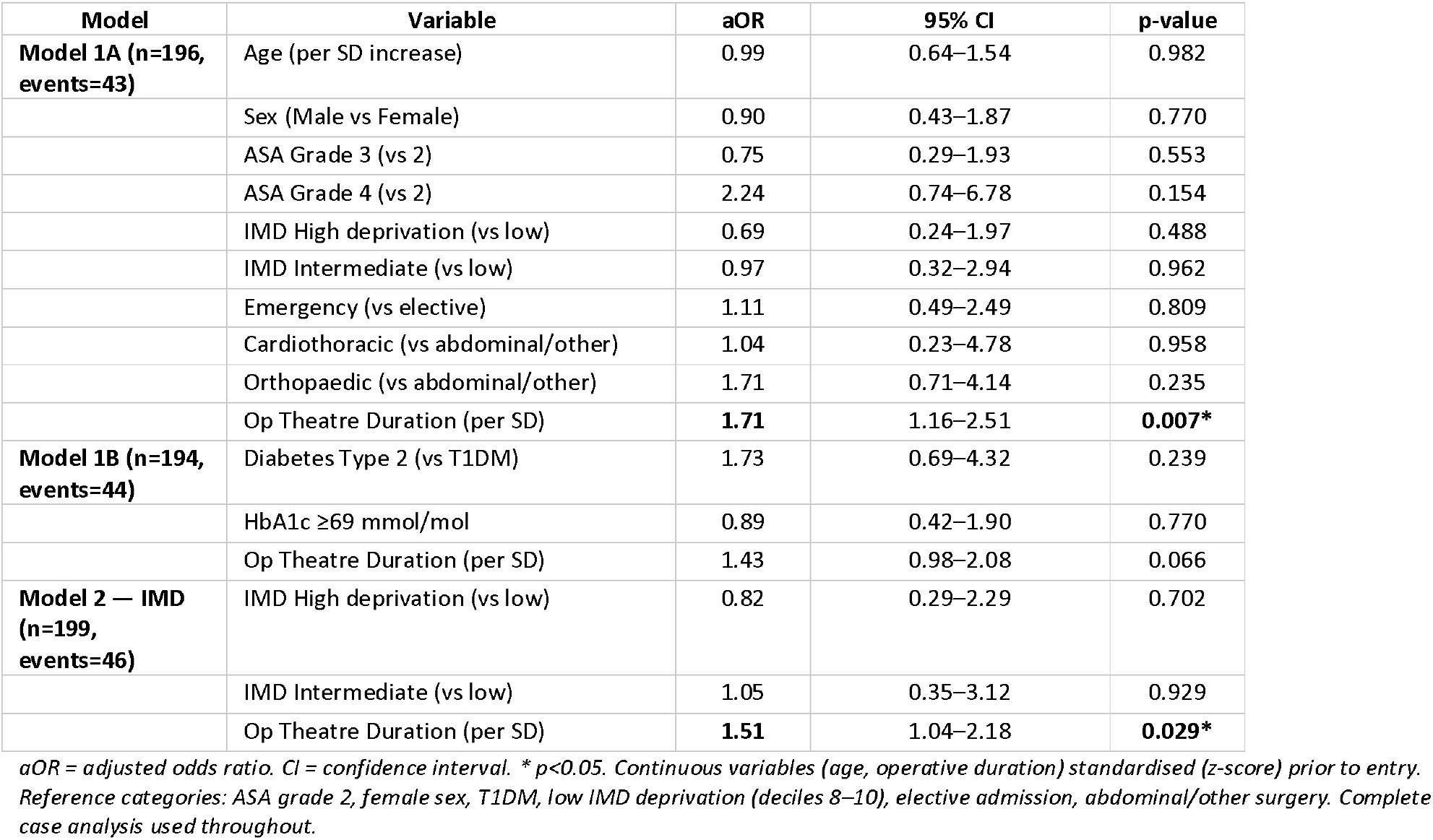
Multivariable Logistic Regression: Adjusted Odds Ratios for Postoperative Complications.

**Figure 2.**
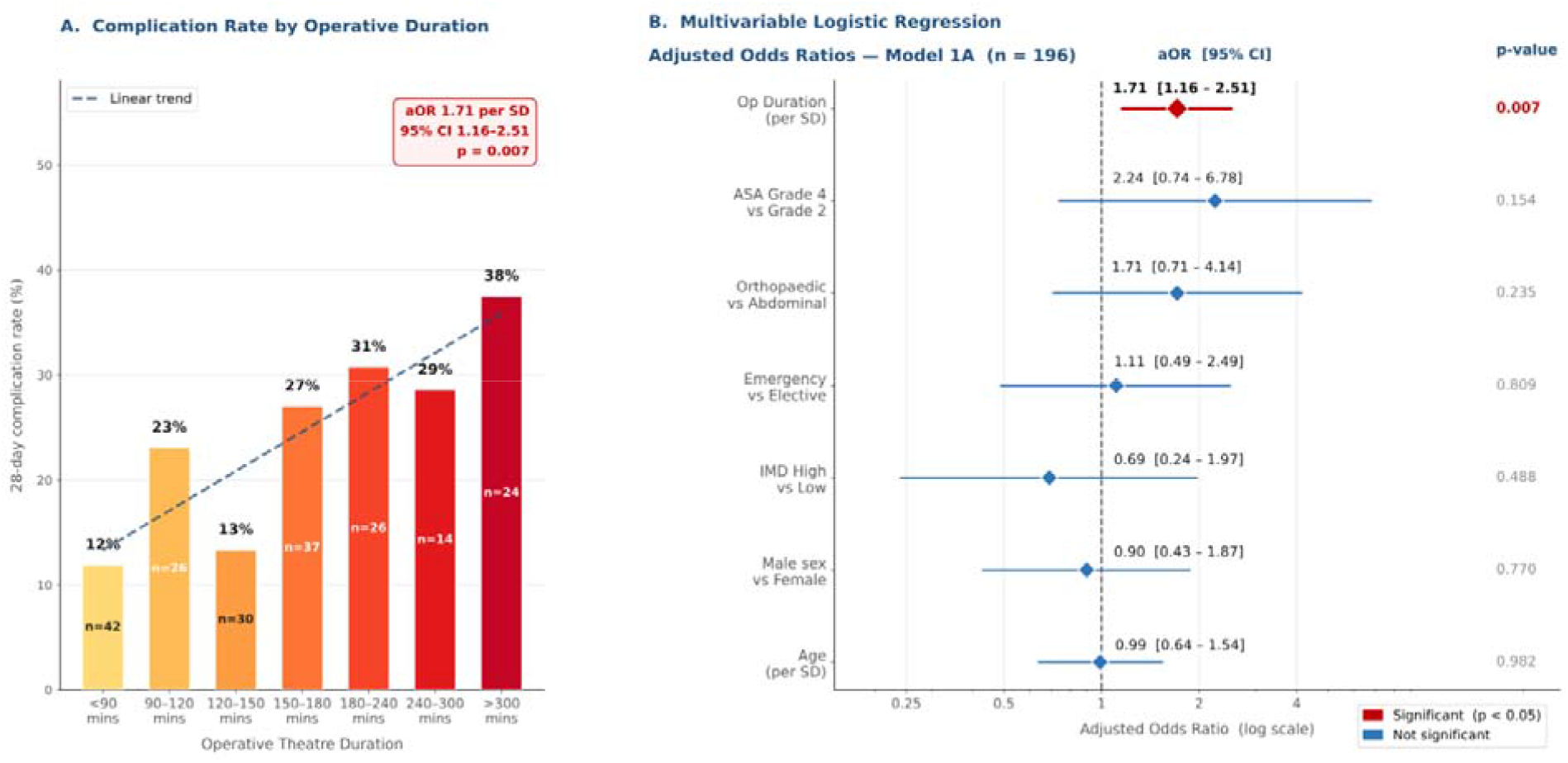
Key Predictors of Postoperative Complications. Panel A: 28-day complication rate (%) within each operative duration bin; n = total patients per bin; dashed line = linear trend (ordinary least squares). Panel B: Forest plot of adjusted odds ratios (aOR) with 95% confidence intervals from multivariable logistic regression Model 1A (n=196 complete cases, 43 events). Log scale. Red = statistically significant (p<0.05). Reference: ASA 2, female, T1DM, IMD decile 8–10, elective, abdominal/other surgery.

In Model 1B (n=194, additionally including HbA1c and diabetes type), neither diabetes type (aOR 1.73, p=0.239) nor HbA1c ≥69 mmol/mol (aOR 0.89, p=0.770) was independently associated with complications. The association with operative duration was attenuated to a trend-level significance (aOR 1.43, 95% CI 0.98–2.08, p=0.066), likely reflecting the smaller complete-case sample.

In Model 2, IMD high deprivation-complication association was not independently associated with postoperative complications compared with low deprivation (aOR 0.82, 95% CI 0.29–2.29, p=0.702). Operative theatre duration remained significant in this model (aOR 1.51, 95% CI 1.04–2.18, p=0.029). Full regression results are presented in Table 6, and a forest plot is shown in Figure 2.

## Discussion

This single-centre retrospective cohort study of 267 adult patients with diabetes mellitus undergoing major surgery at a UK tertiary hospital demonstrates a 28-day postoperative complication rate of 23.1%, a mortality rate of 2.1%, and a readmission rate of 8.1%. In a multivariable logistic regression, operative theatre duration was the only independent predictor of postoperative complications (aOR 1.71 per 1-standard-deviation increase, p=0.007). Age, sex, diabetes type, preoperative HbA1c, ASA grade, admission urgency, and socioeconomic deprivation as measured by IMD were not independently associated with 28-day complications. These findings have important implications for perioperative quality improvement.

The independent association between operative duration and postoperative complications is well supported by the existing literature. A systematic review and meta-analysis of 66 studies by Cheng et al. reported that prolonged operative time was associated with a significantly increased risk of complications, with an approximately 14% incremental increase in risk per additional 30 minutes of operating time^11^. This finding is particularly relevant in diabetic patients, as extended operative duration increases the surgical stress response, worsening insulin resistance and driving sustained hyperglycaemia. Prolonged surgery is also associated with hypothermia (which impairs neutrophil function and promotes wound infection), greater cumulative blood loss and transfusion requirements, and prolonged end-organ hypoperfusion. Critically, operative duration is a modifiable factor. Preoperative planning, team familiarity, optimisation of theatre scheduling, and avoidance of procedural delays are all tractable quality improvement targets. Our findings suggest an association of uniformly high-risk diabetic surgical population (62% ASA grade 3, 69% emergency admissions) and reducing operative duration may be a more immediate lever than glycaemic optimisation alone, though both remain important components of best practice.

The absence of an independent association between preoperative HbA1c and postoperative complications in this cohort requires careful contextualisation. Current UK guidelines recommend an HbA1c threshold of <69 mmol/mol for elective surgery, and the median HbA1c in our cohort was 62.0 mmol/mol. The relatively narrow interquartile range (50.8–75.0) suggests that most patients presented with near-guideline glycaemic control, leaving limited scope for an HbA1c effect to manifest. This likely reflects some degree of pre-surgical optimisation within the tertiary centre setting. Additionally, 69% of our cohort were emergency admissions, for whom preoperative glycaemic optimisation is often not feasible regardless of baseline HbA1c. These findings should not be interpreted as evidence that glycaemic control is unimportant. Rather, they reflect the real-world composition of a high-complexity, emergency-heavy surgical population in which the acute perioperative insult, captured by operative duration, may outweigh chronic glycaemic exposure.

The absence of socioeconomic deprivation as an independent predictor of complications aligns with recent large-scale UK evidence. In the SNAP-2:EpiCCS analysis of 18,901 patients across NHS hospitals, Lusby et al. found that after adjusting for ASA grade, comorbidity, and preoperative anaemia, the association between IMD deprivation and postoperative morbidity and mortality was attenuated and no longer independently significant^12^. Pearse et al accompanying editorial argued that deprivation exerts its perioperative effect primarily through clinical mediators, such as a higher comorbidity burden, later presentation, and worse baseline physiology, rather than as an independent biological risk factor^13^. This interpretation is consistent with our finding that 58.1% of our cohort resided in the most deprived IMD deciles, yet complication rates were statistically indistinguishable across deprivation groups. These findings suggest that within the structured perioperative environment of NHS tertiary care, where national guideline-based glycaemic management pathways are applied irrespective of patients’ background, socioeconomic inequalities in the process of care may be partially mitigated. However, deprivation remains critically important as an upstream driver of the comorbidity burden, late emergency presentations (69% of our cohort), and health-related behaviours that contribute to surgical risk and must remain a priority for pre-hospital and community-based intervention.

The preponderance of infective complications (31 events; 67% of the complication group) reflects established mechanisms, as perioperative hyperglycaemia impairs neutrophil oxidative burst and chemotaxis, reducing host defences against bacterial colonisation. Sepsis (n=13) was the single most common specific complication, consistent with national data. AKI (n=15) was the most common non-infective complication, reflecting the synergistic nephrotoxic burden of surgical stress, contrast media, nephrotoxic antibiotic use, and underlying diabetic nephropathy in this population. The endocrine complication rate was low (DKA n=3, severe hypoglycaemia n=2), which may reflect adequate glycaemic monitoring, though underreporting of minor glycaemic excursions not formally coded as complications cannot be excluded. This study has several methodological strengths. It is based on a systematic EPR screen of all consecutive eligible patients over a five-year period at a tertiary centre, minimising selection bias. A broad range of major surgical specialities is represented, and outcomes are defined using standardised criteria with Clavien-Dindo grading. The use of IMD as a validated area-level deprivation measure enables comparison with published UK literature.

Several limitations must be acknowledged. First, the retrospective design relies on the accuracy and completeness of EPR coding; complications that were not formally documented or coded may have been missed, potentially leading to an underestimation of the true complication rate. Second, complication outcome data were missing for 25.5% of records, representing the principal source of potential bias, even though these patients may not have had any complications. If complications were systematically less likely to be recorded in certain patient groups, results could be subject to informative missingness. Third, with 267 patients and 46 outcome events, the study is powered to detect moderate-to-large effects but may be underpowered to identify modest independent associations; wider confidence intervals for subgroup analyses reflect this limitation. Fourth, the frailty score was available for only 51.7% of patients and could not be included in multivariable models. Fifth, diabetes duration, continuous perioperative glucose monitoring data, insulin regimen details, and preoperative medication optimisation records were incompletely captured in the EPR and therefore absent from analyses. Sixth, as a single-centre study at a UK tertiary hospital with a specific demographic catchment, generalisability to district general hospitals or other regions may be limited. Finally, residual confounding by variables not captured in routine EPR data (e.g., nutritional status, preoperative functional capacity, concurrent infection) cannot be excluded.

## Conclusions

These real-world findings directly support the goals of the ACCORD-BC programme, which aims to advance distributed models of diabetes care within complex, multi-level health systems. By demonstrating that standardised perioperative pathways can attenuate the effects of socioeconomic deprivation and chronic glycaemic control while identifying operative duration as a key modifiable lever, the study provides actionable evidence for integrated care pathways that bridge acute surgical services with primary and community diabetes management, ultimately reducing fragmentation and improving outcomes across the entire care continuum. This study’s emphasis on standardised perioperative protocols to attenuate the impact of glycaemic control and socioeconomic factors directly supports the ACCORD-BC programme’s objectives of advancing distributed diabetes care within complex health systems.

In this five-year retrospective cohort of adult patients with diabetes mellitus undergoing major surgery at a UK tertiary centre, 28-day postoperative complications occurred in 23.1% of patients. Operative theatre duration was the only independent predictor of complications on multivariable analysis. Socioeconomic deprivation and preoperative HbA1c were not independently associated with acute postoperative outcomes. These findings support operative duration as a modifiable quality improvement target in diabetic surgical patients. Prospective multicentre studies with more complete outcome data, perioperative glucose profiles, and richer clinical covariates are needed to further delineate the determinants of adverse outcomes in this high-risk population.

### Declaration of generative AI and AI-assisted technologies in the manuscript preparation process

During the preparation of this work, the author(s) used Grammarly for grammatical and spelling correction. The author(s) reviewed and edited the output as needed and take full responsibility for the content of the published article

## Data Availability

All data produced in the present study are available upon reasonable request to the authors

